# Cycling for climate and public health: a missed opportunity for France

**DOI:** 10.1101/2022.04.14.22273838

**Authors:** Emilie Schwarz, Marion Leroutier, Philippe Quirion, Kévin Jean

## Abstract

In addition to its potential contribution to reaching climate targets, cycling may generate substantial population-level health benefits through the physical activity it requires. Due to the lack of nationally representative mobility data, the health impact of current levels of cycling is still unknown for France. Relying on a health impact assessment framework and using recent nationally-representative data on mobility, we assessed the health and related economic benefits of cycling in 2018-2019 in France. We show that such benefits remain moderated and fall short when compared to those estimated in other countries with high cycling levels. We argue that cycling in France did not receive the attention and investments it deserves over the past decade and thus represents a missed opportunity for climate action and public health.

## Introduction

A recent report released by the French Agency for Food, Environmental and Occupational Health & Safety estimates that 95% of the French adult population faces a risk of deteriorating health due to lack of physical activity [1]. The agency highlights the importance, among others, of collective mobility choices in contributing to this situation. Among these, cycling may represent a particularly relevant active transportation option in France, as an estimated 60% of commuting trips shorter than 5 km are still made by car [2]. Moreover, the transportation sector is responsible for 31% of the national greenhouse gas emissions in France, representing the highest emitting sector [3]. With more than half of these emissions attributable to individual motorized vehicles, encouraging a shift to low-carbon modes such as cycling is an important lever to meet national emission reduction targets.

In countries with high cycling levels, it has been assessed that physical activity through cycling generates substantial population-level health benefits [4]. Due to the scarcity of country-level representative mobility data, the contribution of current levels of cycling is still unknown for France. Until recently, the latest nationally-representative mobility survey was conducted in 2008. In early 2022, the data was released from the most recent round of this mobility survey, the *Enquête mobilité des personnes* (*People’s mobility survey*) conducted in 2018-2019 [5]. We used this updated data to assess the health and related economic benefits of cycling in 2018-2019 in metropolitan France.

## Methods

Demographic data consisting of age-specific population sizes and mortality rates were retrieved from the National Institute of Statistics and Economic Studies (INSEE), using the central projection scenario with low fertility.[6]

Mobility data were extracted from the 2019 *Enquête mobilité des personnes* (*People’s mobility survey*). The 2019 round collected data from 13,825 households in metropolitan France.[5] One member of each household was selected and questioned about their trips across the previous day. The sampling design and sampling weights ensure that the survey data is nationally representative Of travel behaviours for individuals aged 6 years or older, across weekdays and weekends. Distances travelled were calculated based on reported departures and arrival location, and converted into times considering an average speed of 14.9 km.h^1^ [7].

Based on the approach described by Fishman et al. (2015), we used age-specific weekly cycling minutes, population counts, and mortality rates for 2019, and assumed a linear dose-response function of 10% reduction in all-cause mortality for 100 minutes of cycling per week [8]. Given that the meta-analysis by Kelly et al considered all-cause mortality, any increase in mortality in terms of road accidents and exposure to air pollution is already accounted for in the dose-response estimate. Risk reduction estimates were scaled at the yearly level assuming a linear dose-response function. The age range considered for a reduction of mortality attributable to physical activity increase was 20-89 years. Younger and older ages were disregarded because the evidence for the health effects of physical activity in those age categories are small.

We estimated the yearly number of premature deaths averted by active transportation for each age. We also calculated the years of life lost (YLL) averted by active transportation, along with the annual monetary benefits of cycling based on the standard value of statistical life year (VSLY). National guidelines for the socioeconomic evaluation of public investments recommended a VSLY of €135,000 in 2019 [9]. Lastly, we used age-specific death rates to estimate the increase in life expectancy due to cycling at 2019 levels.

## Results

The average weekly minutes of cycling per person in 2019 in France was estimated at 8.7 min.w^-1^, peaking at 13.4 min.w^1^ among the 70-74-year-old group (Table 1). Overall, we estimate that 2019 levels of cycling represented nearly 2,900 yearly premature deaths and 38 000 YLL prevented. This translates into a <1-month increase of life expectancy in the general population. We evaluated that the annual monetary benefits of cycling reached €5 billion in France.

**Table 1:** Health impact assessment of 2019 levels of cycling in metropolitan France among adults aged 20 to 89 years.

| Age group, years | Average weekly minutes of cycling per person<sup>1</sup> | Population, x1,000<sup>2</sup> | Mortality rate, per 100,000<sup>2</sup> | Mortality rate reduction<sup>3</sup>, % | Annual # of premature death prevented | Annual benefits of cycling, in € million<sup>4</sup> | Increase in life expectancy, in months |
| --- | --- | --- | --- | --- | --- | --- | --- |
| 20-24 | 4.9 | 3811 | 38 | 0.49 | 7 | 58 | 0.007 |
| 25-29 | 7.1 | 3927 | 42 | 0.71 | 12 | 87 | 0.010 |
| 30-34 | 10.2 | 4128 | 55 | 1.02 | 23 | 157 | 0.017 |
| 35-39 | 11.1 | 4113 | 87 | 1.11 | 40 | 242 | 0.026 |
| 40-44 | 11.0 | 3939 | 145 | 1.10 | 62 | 340 | 0.039 |
| 45-49 | 11.4 | 4374 | 238 | 1.14 | 118 | 563 | 0.058 |
| 50-54 | 7.1 | 4297 | 359 | 0.71 | 110 | 451 | 0.047 |
| 55-59 | 5.8 | 4187 | 484 | 0.58 | 117 | 402 | 0.044 |
| 60-64 | 8.9 | 3968 | 664 | 0.89 | 234 | 644 | 0.076 |
| 65-69 | 9.2 | 3838 | 946 | 0.92 | 335 | 695 | 0.090 |
| 70-74 | 13.4 | 3202 | 1449 | 1.34 | 622 | 886 | 0.153 |
| 75-79 | 9.7 | 2109 | 2468 | 0.97 | 505 | 367 | 0.128 |
| 80-84 | 5.3 | 1808 | 4374 | 0.53 | 420 | 71 | 0.075 |
| 85-89 | 2.5 | 1336 | 8099 | 0.25 | 269 | 166 | 0.032 |
| **Total or average** | **8.7** | **49037** | **834** | **0.87** | **2875** | **5129** | **0.802** |

<sup>1</sup>Based on reported trips of the day preceding participants' interviews for the *Enquête mobilité des* *personnes (People's mobility survey)* [5]. Distances travelled were calculated based on reported departures and arrival places, and converted into times considering an average speed of 14.9 km.h<sup>-1</sup> [7].
<sup>2</sup>Retrieved from the National Institute of Statistics and Economic Studies (INSEE), 2007-2060 population projections.
<sup>3</sup>Assuming a 10% reduction in all-cause mortality for 100 minutes of cycling per week with a linear dose-response function [8].
<sup>4</sup>Based on the standard value of a statistical life year of € 135,000 (in 2019€), as per national recommendations for the socioeconomic evaluation of public investments (publicly available here).

## Discussion

The French health benefits of cycling appear low when compared to those reported for the Netherlands back in 2010-2013, where cycling contributed to a half-a-year-longer life expectancy and annual monetary benefits of cycling reached €18.6 billion, for a population about 4 times smaller than France [4]. However, it is worth highlighting that even if they appear moderate, the French levels of cycling already provide substantial monetary benefits which would further increase with an augmentation in bike practice. Hence, public investments in cycling-promotion policies, such as improved cycling infrastructures and facilities, are likely to yield a high cost-benefit ratio in the medium term.

Despite repeated calls to decrease motorized trips for the interest of the climate and to increase active mobility for public health, the level of cycling for France did not evolve substantially between 2008 and 2019. The modal share of cycling remained the same (2.7%) across both survey rounds [5]. Therefore, we argue that public policies to promote cycling represent a missed opportunity for both climate targets and public health in France over the last decade.

Unfortunately, recent announces do not reveal a massive shift in mobility policies. As in many other European countries, the French government has recently cut taxes by 15 cents per litter of gasoline in response to the latest increase in fuel prices [10]. This policy will cost about €2 billion over 4 months. In comparison, the 2018 French strategy to promote cycling, one of the most ambitious ever implemented in the country, relied on public investments of only €0.45 billion over 7 years [11].

We hope that the health and climate benefits of cycling will soon be recognized and that it will receive the attention and investments it deserves. Recently, due to the Covid-19 impact on public transportation, some local authorities have rapidly incentivized cycling by rolling out pop-up bike lanes. This resulted in a large short-term increase in cycling, including in France [12]. Commitment of national and local authorities is critical to sustaining these changes and the contribution of cycling to public and planetary health.

## Data Availability

All data produced in the present work are contained in the manuscript.

https://www.statistiques.developpement-durable.gouv.fr/resultats-detailles-de-lenquete-mobilite-des-personnes-de-2019

## Notes

<u>Conflict of interest</u> The authors declare no conflict of interest.

### Competing Interest Statement

The authors have declared no competing interest.

### Funding Statement

This study did not receive any funding

